# Investigation on the current situation of stroke patients in northernern Henan Province of China

**DOI:** 10.1101/2021.07.04.21259998

**Authors:** Zhenhua Li, Qingcheng Yang, Xiangdong Zhang, Yanping Guo, Jiangang Zhang, Ke Sun, Junzheng Yang

**Affiliations:** Department of Neurosurgery, Anyang People’s Hospital, Anyang, Henan, 455001, China

**Author notes:** Correspondence to Junzheng Yang, Department of Neurosurgery, Anyang People’s Hospital, Anyang, Henan, 455001, China.

**Keywords:** stroke, prevention, investigation, northern Henan Province

## Abstract

**Objectives:** To analyze the basic situations and clinical characteristics of stroke patients in our hospital from May 2018 to April 2021, lay a foundation for the prevention and reasonable treatment of stroke patients in northern Henan Province.

**Methods:** The basic information of 835 stroke patients in our hospital was collected and classified according to age, gender, bad habits, accompanied diseases and drug use before admission to hospital, and severity of the stroke patients were also evaluated according to mRS scoring standard.

**Results:** A total of 835 stroke patients were collected from May 2018 to April 2021 in our hospital. The age range of stroke patients was 28-95 years old, 96.29% stroke patients was above 40 years old; there were 202 stroke patients with smoking history and 225 stroke patients with drinking history; Among the 835 stroke patients, hypertension, cerebral infarction and diabetes mellitus were the main accompanied diseases. Antihypertensive drugs (506 cases), antiplatelet drugs (208 cases), statins (173 cases) and antidiabetic drugs (143 cases) were the main therapeutic drugs in stroke patients before admission in the northern Henan Province; the results of mRS scoring standard showed that among 835 stroke patients, there were 609 cases with milder symptoms, accounting for 82.84% (there were 330 stroke patients with 1 points, 279 stroke patients with 2 points, and 83 stroke patients with 3 points), and 120 cases with severe symptoms, accounting for 14.37% (55 cases with 4 points, 65 cases with 5 points).

**Conclusion:** The age of stroke patients in northern Henan Province was mainly over 40 years old, most of stroke patients were in the early stage of stroke; smoking, drinking, hypertension and diabetes mellitus were main risk factors of stroke. And there was a sex difference between male stroke patients and female stroke patients in stroke risk factors smoking and hypertension. those data may help us for active prevention and rational drug use for stroke in clinic.

## 1 Introduction

Statistical studies have found that stroke has become the third largest diseases in recent years, which has the highest mortality rate and is becoming the main cause of death in 27 of 33 provinces in China ^[1, 2, 3]^. It was reported that there are 2.4 million new stroke patients increasing every year, and 75% of stroke patients were accompanied with different degrees of disability caused by stroke^[4, 5]^; stroke could lead to subsequent vascular cognitive impairment and increased the risk of many other diseases, such as Alzheimer’s disease; stroke also cause great losses to individuals and families, both in spirit and economy^[6,7]^.

More than 80% of stroke can be prevented, stroke prevention can not only prolong life span, but also ensure the quality of life and cognitive function. Early detection of stroke symptoms and understanding of the basic situation of stroke patients are of great significance to the prevention and treatment of stroke, For this reasons, this article investigated and summarized the age, gender, bad habits, accompanying diseases, drug use and stroke severity of stroke patients before admission to our hospital from May 2018 to April 2021. The article was reported as follows.

## 2 Data and Methods

### 2.1 Investigation subjects

835 stroke patients were treated in our hospital from May 2018 to April 2021.

### 2.2 Investigation methods

835 stroke patients treated in our hospital from May 2018 to April 2021 were classified according to their age, gender, bad habits, accompanied diseases and drug use before admission to the hospital; 835 stroke patients were also scored by mRS scoring standard before admission.

### 2.3 Admission criteria for stroke patients

Based on the World Health Organization (WHO) criteria, stroke was defined as “rapidly developing clinical syndrome of focal or global abnormalities of cerebral function, lasting more than 24 hours or leading to death, with no apparent cause other than that of vascular origin”^[8]^. Any nervous system abnormalities induced by trauma, metabolic disorder, tumor, or central nervous system infections were excluded.

### 2.4 mRS scoring standard

According to the standard and the severity of the patient’s condition, it can be divided stroke into six grades: 0 points: totally asymptomatic. Although there may be slight symptoms, the patient has not noticed of any new functional limitation and symptoms since stroke. 1 points: no obvious disability was found in spite of symptoms; be able to complete all the duties and activities that often engage in. 2 points: mild disability; unable to complete all the previous activities, but able to handle personal affairs without help. 3 points: moderate disability; need some help, but don’t need help to walk. 4 points: severe disability; unable to walk without the help of others, and unable to take care of their own physical needs. 5 points: severe disability; bedridden, incontinence, continuous care and care are required. Although trained nurses are not required, they need to be looked after several times during the day and night.

## 3 Results

### 3.1 General information

Of the 835 stroke patients were treated in our hospital from May 2018 to April 2021, there were 565 male patients, accounting for 67.66%; There were 270 female patients, accounting for 32.34%; the age range of the 835 stroke patients was 28-95 years old; 96.29% of stroke patients was above 40 years old. With the increase of age, the proportion of stroke patients increased, and the number of male stroke patients was higher than that of female stroke patients (Table 1).

**Table 1.** Age distribution and sex distribution of 835 stroke patients in the northern Henan Province

| Age | Male | Female |
| --- | --- | --- |
| <30 | 3 (0.36%) | 0 |
| 31-40 | 18 (2.16%) | 10 (1.20%) |
| 41-50 | 57 (6.83%) | 32 (3.83%) |
| 51-60 | 120 (14.37%) | 65 (7.78%) |
| 61-70 | 199 (23.83%) | 90 (10.78%) |
| >71 | 168 (20.12%) | 73 (8.74%) |
| Total | 565 (67.66%) | 270 (32.34%) |

Among the 835 stroke patients, 202 cases had smoking history, there were 190 male (22.75%); 112 female (13.41%), and the difference between male patients and female patients with smoking history was statistically significant (P<0.05); There were 225 patients with drinking history, including 153 males (68%); 72 females (32%), and there was no statistical significance between male patients and female patients with drinking history (P>0.05) (Table 2).

**Table 2.**
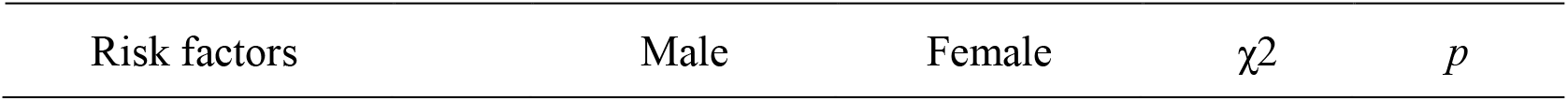
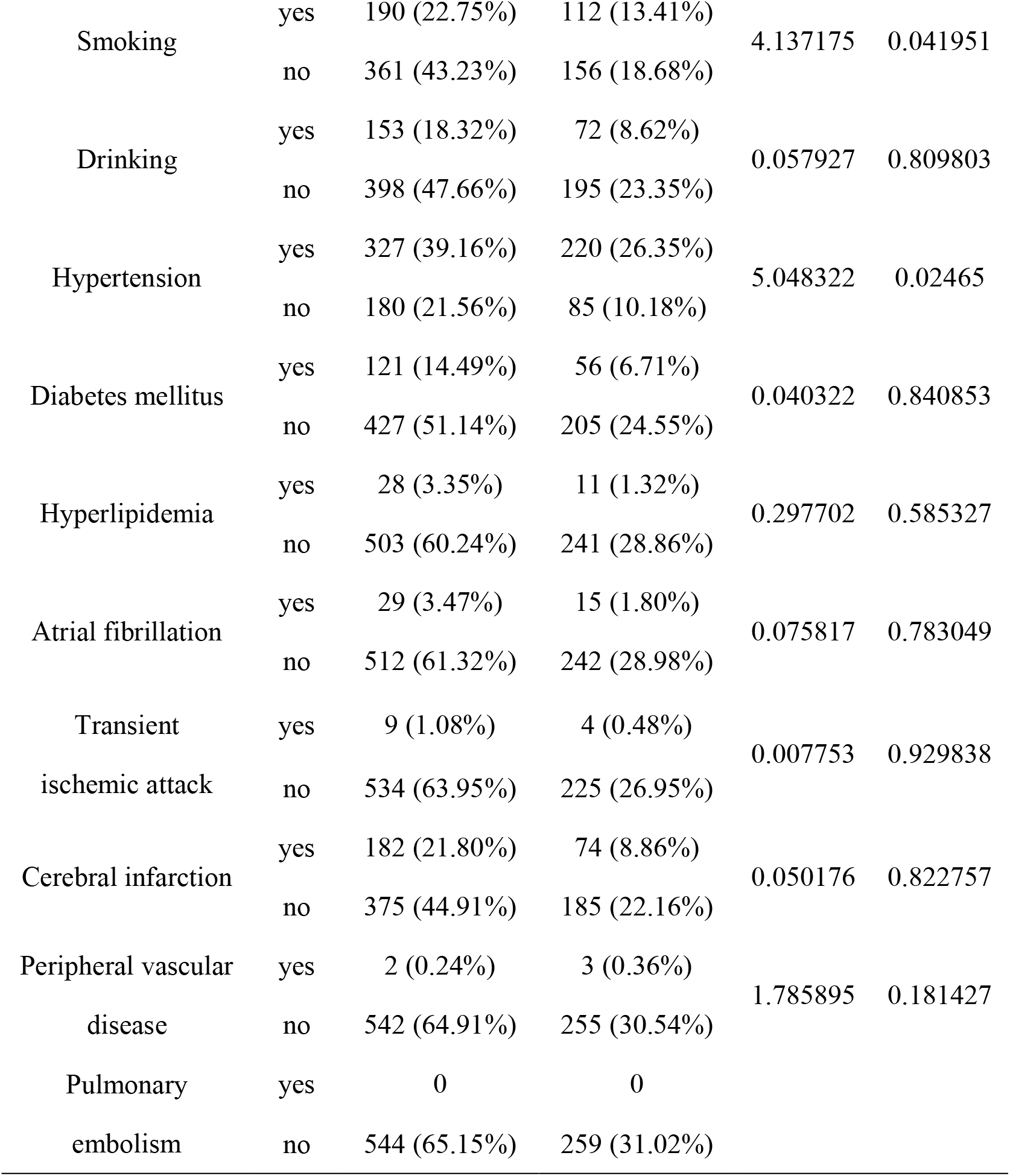
Smoking, drinking and common accompanied diseases of 835 stroke patients in the northern Henan Province

| Risk factors | Male | Female | $\chi^2$ | $p$ |
| --- | --- | --- | --- | --- |

|  |  |  |  |  |  |
| --- | --- | --- | --- | --- | --- |
| Smoking | yes | 190 (22.75%) | 112 (13.41%) | 4.137175 | 0.041951 |
|  | no | 361 (43.23%) | 156 (18.68%) |  |  |
| Drinking | yes | 153 (18.32%) | 72 (8.62%) | 0.057927 | 0.809803 |
|  | no | 398 (47.66%) | 195 (23.35%) |  |  |
| Hypertension | yes | 327 (39.16%) | 220 (26.35%) | 5.048322 | 0.02465 |
|  | no | 180 (21.56%) | 85 (10.18%) |  |  |
| Diabetes mellitus | yes | 121 (14.49%) | 56 (6.71%) | 0.040322 | 0.840853 |
|  | no | 427 (51.14%) | 205 (24.55%) |  |  |
| Hyperlipidemia | yes | 28 (3.35%) | 11 (1.32%) | 0.297702 | 0.585327 |
|  | no | 503 (60.24%) | 241 (28.86%) |  |  |
| Atrial fibrillation | yes | 29 (3.47%) | 15 (1.80%) | 0.075817 | 0.783049 |
|  | no | 512 (61.32%) | 242 (28.98%) |  |  |
| Transient ischemic attack | yes | 9 (1.08%) | 4 (0.48%) | 0.007753 | 0.929838 |
|  | no | 534 (63.95%) | 225 (26.95%) |  |  |
| Cerebral infarction | yes | 182 (21.80%) | 74 (8.86%) | 0.050176 | 0.822757 |
|  | no | 375 (44.91%) | 185 (22.16%) |  |  |
| Peripheral vascular disease | yes | 2 (0.24%) | 3 (0.36%) | 1.785895 | 0.181427 |
|  | no | 542 (64.91%) | 255 (30.54%) |  |  |
| Pulmonary embolism | yes | 0 | 0 |  |  |
|  | no | 544 (65.15%) | 259 (31.02%) |  |  |

### 3.2 Accompanying diseases

Among the 835 stroke patients, 547 patients were accompanied with hypertension, including 327 males (39.16%) and 220 females (26.35%); 256 patients accompanied with cerebral infarction, 176 patients accompanied with diabetes mellitus, 44 patients accompanied with atrial fibrillation, 39 patients accompanied with metabolic disorders, 13 patients accompanied with transient ischemic attack, 5 patients accompanied with peripheral vascular disease, and there was no statistically significant difference between male and female patients in all the accompanied diseases (p>0.05) (Table 2).

### 3.3 Drug use of stroke patients before admission

506 stroke patients used antihypertensive drugs, including 339 male patients, accounting for 41.80%, 167 female patients, accounting for 20.59%; Secondly, 203 stroke patients used antiplatelet drugs, including 128 male patients (15.92%) and 75 female patients (9.33%); there were 173 patients using statins, including 113 male patients (14.04%) and 60 female patients (7.45%); There were 143 patients used antidiabetic drugs, 58 patients used Chinese patent medicine and 19 patients used anticoagulant drugs. There was no statistical significance between male and female patients (P>0.05) (Table 3).

**Table 3.** Therapeutic drugs use of 835 stroke patients in the northern Henan Province

| Drug type | Drug use | Male | Female | $\chi^2$ | $p$ |
| --- | --- | --- | --- | --- | --- |
| Antihypertensive drugs | yes | 339 (41.80%) | 167 (20.59%) | 0.202904 | 0.652387 |
|  | no | 209 (25.77%) | 96 (11.84%) |  |  |
| Antiplatelet drugs | yes | 128 (15.92%) | 75 (9.33%) | 2.34862 | 0.125394 |
|  | no | 414 (51.49%) | 187 (23.26%) |  |  |
| Statins | yes | 113 (14.04%) | 60 (7.45%) | 0.457734 | 0.498685 |
|  | no | 430 (53.42%) | 202 (25.09%) |  |  |
| Antidiabetic drugs | yes | 92 (11.57%) | 51 (6.42%) | 0.755854 | 0.384629 |
|  | no | 444 (55.85%) | 208 (26.16%) |  |  |
| Chinese Medicine<br>(cerebrovascular disease<br>related) | yes | 39 (5.05%) | 17 (2.20%) | 0.154344 | 0.694418 |
|  | no | 481 (62.25%) | 236 (30.53%) |  |  |
| Anticoagulants | yes | 13 (1.65%) | 6 (0.76%) | 0.009495 | 0.922376 |
|  | no | 518 (65.74%) | 251 (31.85%) |  |  |

### 3.4 Evaluation of Severity of stroke patients before admission

The mRS score of 835 stroke patients showed that there was no completely asymptomatic patient in 835 stroke patients; The number of patients with 1 points was 330, including 239 males and 91 females; The number of patients with 2 points was 279, including 193 males and 86 females; The number of patients with 3 points was 83, including 42 males and 42 females; The number of patients with 4 points was 55, including 33 males and 22 females; The number of patients with 5 points was 65, including 42 males and 23 females. The population of male stroke patients was higher than the population of female stroke patients in all five severity degree (Table 4).

**Table 4.** mRS score before admission in 835 stroke patients

| Severity of stroke | Cases |  |  |
| --- | --- | --- | --- |
|  | Male | Female | Total |
| 0. No symptoms | 0 | 0 | 0 |
| 1. Although there are symptoms, there is no obvious dysfunction and can complete all daily duties and activities | 239(28.62%) | 91(10.90%) | 330(39.52%) |
| 2. Slightly disabled, unable to complete all activities before illness, but able to take care of their own affairs without help | 193(23.11%) | 86(10.30%) | 279(33.41%) |
| 3. Moderate disability, need some help, but walking without help | 42(5.03%) | 41(4.91%) | 83(9.94%) |
| 4. Severely disabled, unable to walk independently, unable to meet their own needs without the help of others | 33(3.95%) | 22(2.63%) | 55(6.59%) |
| 5. Severe disability, bedridden, incontinence, requiring continuous care and attention | 42(5.03%) | 23(2.75%) | 65(7.78%) |

## 4 Discussion

Many articles have been reported that there was a sex difference in stroke incidence and mortality, usually incidence of stroke in women was lower than in men^[9, 10, 11]^. In our article, 835 stroke patients were treated in our hospital from May 2018 to April 2021, 565 stroke patients were male, accounting for 67.66%; there were 270 female stroke patients, accounting for 32.34%; the number of male stroke patients was significantly higher than that of female stroke patients (67.66 versus 32.34%), and the difference became more and more obvious with the increase of age; the largest difference was found in patients aged 61-70 years old (male 23.83% versus female 10.78%) or above 71 years (male 20.12% versus female 8.14%), which may be related to the bad living habits of men such as drinking and smoking; we also found that the age of stroke patients is mainly above 40 years old, but it should be noted that among the male stroke patients, there were 3 male stroke patients under 30 years old, which may indicate that the age of stroke patients in Northern Henan tends to the young.

Stroke is a cerebrovascular disease mainly caused by atherosclerosis. Hypertension, diabetes mellitus, smoking and drinking can often cause the aggravation and rapid growth of stroke patients. Hypertension is considered to be the most important risk factor for all vascular diseases including stroke. Statistical studies have found that the relative risk coefficient of hypertension and stroke is as high as 5.13, smoking and drinking increased the risk of stroke by 1.8 times^[12, 13]^. In this study, hypertension, cerebral infarction and diabetes are the three major accompanied diseases of stroke in the northern of Henan Province. Among them, there were 547 cases of stroke with hypertension, male patients (327 cases, 39.16%) and female patients (220 cases, 26.35%), the difference was statistically significant between male patients and female patients (*P*<0.05), indicating that there was not only a relationship between hypertension and stroke, but also a sex difference, which needs to be paid attention to by clinicians. Among 835 stroke patients, 225 patients had a drinking history, accounting for 26.94%; there were 202 patients with smoking history, accounting for 24.19%, indicating that the bad living habits of stroke patients maybe also key risk factors of stroke in northern Henan Province.

Among the 835 cases of stroke patients were treated in our hospital, 506 stroke patients used antihypertensive drugs for stroke treatment, demonstrated that hypertension is still the main accompanied diseases of stroke in northern Henan Province. Then we evaluated 835 stroke patients with mRS scoring standard, found that 509 stroke patients were 1 points or 2 points, 83 stroke patients were 3 points, and 120 stroke patients were 4 points or 5 points; results showed that most stroke patients in northern Henan Province were in milder symptoms of stroke (75.69%), but there were 203 patients with moderate and severe symptoms, accounting for 24.31%; stroke patients will give priority to oral medication for prevention and treatment. The reason for this situation may be related to the economic situation in northern Henan and the education level of patients.

## 5 Conclusions

The age, gender, bad habits, accompanied diseases and drug use of 835 stroke patients in northern Henan Province were collected and classified; the mRS scoring standard for evaluation of 835 patients with stroke before admission was conducted. It was found that the age of stroke patients in northern Henan province was mainly over 40 years old, most of them are in the early stage of the stroke; and smoking, drinking, hypertension and diabetes mellitus were main risk factors of stroke. Antihypertensive drugs, antiplatelet drugs, statins and antidiabetic drugs were mainly used; population of male stroke patients was higher than female stroke patients, and there was a sex difference between male stroke patients and female stroke patients in stroke risk factors smoking and hypertension. Those evidences may help us for prevention and rational treatment for stroke patients.

## Data Availability

The data used to support the findings of this study are available from the corresponding author upon request.

## 6. Acknowledgements

None.

